# Dynamics of antibodies to SARS-CoV-2 in convalescent plasma donors

**DOI:** 10.1101/2021.01.06.20249035

**Authors:** Maurice Steenhuis, Gerard van Mierlo, Ninotska I.L. Derksen, Pleuni Ooijevaar-de Heer, Simone Kruithof, Floris L. Loeff, Lea C. Berkhout, Federica Linty, Chantal Reusken, Johan Reimerink, Boris Hogema, Hans Zaaijer, Leo van de Watering, Francis Swaneveld, Marit J. van Gils, Berend Jan Bosch, Marieke van Ham, Anja ten Brinke, Gestur Vidarsson, Ellen C. van der Schoot, Theo Rispens

## Abstract

The novel SARS-CoV-2 virus emerged in late 2019 and has caused a global health and economic crisis. The characterization of the human antibody response to SARS-CoV-2 infection is vital for serosurveillance purposes as well for treatment options such as transfusion with convalescent plasma or immunoglobin products derived from convalescent plasma. In this study, we measured antibody responses in 844 longitudinal samples from 151 RT-PCR positive SARS-CoV-2 convalescent adults during the first 34 weeks after onset of symptoms. All donors were seropositive at the first sampling moment and only one donor seroreverted during follow-up analysis. Anti-RBD IgG and anti-nucleocapsid IgG levels slowly declined with median half-life’s of 62 and 59 days during 2-5 months after symptom onset, respectively. The rate of decline of antibody levels diminished during extended follow-up. In addition, the magnitude of the IgG response correlated with neutralization capacity measured in a classic plaque reduction assay as well in our in-house developed competition assay. The result of this study gives valuable insight into the longitudinal response of antibodies to SARS-CoV-2.

## Introduction

The novel coronavirus, severe acute respiratory syndrome coronavirus 2 (SARS-CoV-2), the causative agent of the on-going coronavirus disease (COVID-19) pandemic emerged in Wuhan (China) in December 2019. SARS-CoV-2 is classified under the *Betacoronavirus* 2B and is closely related to SARS-CoV (>80% genomic similarity) and MERS-CoV (50% genomic similarity), which are known of previous outbreaks [1, 2]. COVID-19 is a disease that is associated with a wide spectrum of disease severity, ranging from asymptomatic to acute respiratory distress syndrome and is already responsible for more than 1 million deaths worldwide [3].

Besides vaccination, prevention of infection with SARS-CoV-2 might be achieved by transfusion with COVID-19 convalescent plasma (CCP), plasma collected from individuals after recovery from COVID-19, or immunoglobulin products derived from CCP [4-6]. This therapy would be especially relevant for immunocompromised individuals. Although it has been shown that CCP therapy is safe [7] and it has been approved by the FDA for treatment, the clinical effect especially in severely ill patients seems to be limited [8]. The most potent CCP units are in theory those containing the highest amounts of neutralizing antibodies against SARS-CoV-2. To select convalescent plasma donors with high neutralizing antibody titers it is important to understand the dynamics of antibodies against SARS-CoV-2 in the period after recovery from SARS-CoV-2.

Virtually all PCR-confirmed patients develop IgM, IgA and IgG antibodies against the virally encoded surface glycoproteins spike (S) and nucleocapsid protein (NP) [9-11]. The S protein mediates binding of the virus particle to angiotension converting enzyme-2 (ACE2) on target cells through its receptor binding domain (RBD) [12, 13], enabling merging of viral and target cell membrane. While a large fraction of anti-S antibodies are directed against RBD, of which many are neutralizing [14, 15], anti-S antibodies binding outside of the RBD may also contribute to neutralization [14, 15]. In addition, antibody levels seem to vary depending on the infection duration and severity of disease, and anti-S and anti-NP antibodies correlate well [16].

Seroprevalence studies have demonstrated that antibodies to SARS-CoV-2 can be detected up to at least 3-8 months after disease recovery [17-20]. However, seroprevalence depends on the characteristics of the study population, on the detection methods used and do not report on the quantitative aspects of the antibody response. Many short-term studies consistently show a seroconversion of IgG, IgA and IgM antibodies against the viral proteins S and NP within 1-3 weeks after symptom onset, depending on disease severity [9-11, 21]. Less information is available about the long-term course of antibody titers. Several studies investigated the longitudinal antibody response and found that 1 month after onset of symptoms antibody levels reach a plateau followed by rapidly declining IgM and IgA titers, whereas IgG titers seems to remain high up to 6 months [18, 19, 22-26]. A recent study by Dan *et al*., in which patients were followed up to 8 months, consistently showed only a modest decline in anti-S IgG titers as well as neutralizing antibody titers [20]. Limitations of these studies include a sometimes limited cohort size, and in particular the number of longitudinal data points available for each subject, which restricts the possibilities to analyze trends in antibody levels over time.

Here we collected samples from 151 RT-PCR positive SARS-CoV-2 recovered adults donating convalescent plasma over a study period of up to 34 weeks. The median number of samples per donor was 5 (IQR 4-7; range 2-18), which allowed a more detailed analysis of individual trends over time.

## Materials and Methods

### Samples and donors

Plasma samples were obtained from RT-PCR positive SARS-CoV-2 recovered adult individuals (n=151) donating convalescent plasma at Sanquin Blood Bank (Amsterdam, the Netherlands) who enrolled the CP program of Sanquin between March 30, 2020 and September 6, 2020. Donors were included if tested positive in a SARS-CoV-2 PCR, which during this period was only provided to individuals displaying COVID-19 related symptoms, and were symptom-free for at least two weeks. Details are provided in Table 1. Inclusion was contingent upon the availability of at least two samples with 30 days in between and samples were collected between March 30 and August 14, 2020 (period 1), yielding 694 samples. Additional samples were collected based on availability for 55 donors up to November 11, 2020 (period 2), yielding another 150 samples. During this period many donors dropped out due to insufficient titers. Some donors with low titers continued as regular plasma donor. The median number of samples per donor was 5 (IQR 4-7; range 2-18).

**Table 1.** Baseline table for the convalescent plasma donors used in this study.

|  | Tot<br>(n = 151) | Male<br>(n = 105) | Female<br>(n = 46) | *p* | Non-Hosp.<br>(n = 128) | Hosp.<br>(n = 23) | *p* |
| --- | --- | --- | --- | --- | --- | --- | --- |
| age (yr) | 45 (35-55)<sup>a)</sup> | 48 (36-55) | 42 (29-51) | 0.058 | 42 (31-53) | 54 (48-58) | <0.001 |
| M/F (%) | 70 |  |  |  | 64 | 100 | <0.001 |
| Non-hosp./hosp. (%) | 85 | 78 | 100 | <0.001 |  |  |  |
| first donation<br>(days after<br>symptom onset) | 59 (47-74) | 59 (48-74) | 54 (38-80) | 0.61 | 54 (45-74) | 63 (52-76) | 0.13 |
| IgG-RBD<br>(AU/mL)<sup>b)</sup> | 24 (9-70) | 30 (10-89) | 17 (9-28) | 0.009 | 18 (8-39) | 113 (70-223) | <0.001 |
| IgG-NP<br>(AU/mL)<sup>b)</sup> | 28 (8-92) | 35 (10-105) | 16 (6-56) | 0.012 | 19 (7-66) | 117 (45-300) | <0.001 |
| IgM-RBD (%)<sup>b)</sup> | 49 | 55 | 35 | 0.024 | 45 | 70 | 0.027 |
| IgA-RBD (%)<sup>b)</sup> | 65 | 70 | 55 | 0.075 | 60 | 91 | 0.004 |
| half-life RBD-IgG<br>(days)<sup>c)</sup> | 62 (48-87) | 59 (47-84) | 69 (53-99) | 0.027 | 64 (49-93) | 54 (47-68) | 0.053 |
| half-life NP-IgG<br>(days)<sup>c)</sup> | 59 (44-82) | 58 (43-78) | 67 (50-108) | 0.023 | 60 (45-84) | 54 (42-64) | 0.12 |

a) numbers between parentheses indicate interquartile range
b) value at first donation
c) obtained from regression analysis

Data and samples were collected only from voluntary, non-remunerated, adult donors as described previously [27], and who provided written informed consent as part of routine donor selection and blood collection procedures, that were approved by the Ethics Advisory Council of Sanquin Blood Supply Foundation. This study has been conducted in accordance with the ethical principles set out in the declaration of Helsinki and all participants provided written informed consent.

### Total and isotype-specific antibody ELISA

Total antibody, IgM and IgA to RBD was measured as described previously [9]. IgG to RBD and NP were measured essentially as described before [9], but with several modifications. RBD and NP proteins were produced as described in Vogelzang *et al*., [9]. IgG1 and IgG3 subclasses were detected by adding 0.5 µg/mL HRP-conjugated monoclonal mouse antihuman IgG1 (MH161.1, Sanquin) or 1.0 µg/mL antihuman IgG3 (MH163.1, Sanquin) diluted in PBS supplemented with 0.02% polysorbate-20 and 0.3% gelatin) (PTG) for 30 minutes. Samples were tested 1:1200 diluted for IgG and IgG1 to RBD, and 1:100 diluted for IgG3 to RBD. A cut-off was based on the 98 percentile of signals of 240 pre-outbreak plasma samples. Signals were quantified using a serially diluted calibrator consisting of pooled convalescent plasma that was included on each plate. This calibrator was arbitrarily assigned a value of 100 AU/mL. For anti-RBD IgG1 and IgG3, the calibrator consisted of recombinantly expressed IgG1 and IgG3 monoclonal antibody. Both antibodies had the same variable regions (clone 1-18), but were engineered with different heavy chains [15, 28]. Results were expressed as arbitrary units per mL (AU/mL) and represent a semi-quantitative measure of the concentrations of IgG antibodies.

### Competition ELISA

Neutralizing capacity of SARS-CoV-2 antibodies was assessed using a competition assay. Serum samples were incubated 125-fold diluted for 60 minutes with 5 ng/mL biotinylated RBD in PTG. Next, 100 uL aliquots were transferred to MaxiSORP microtiter plates (ThermoFisherScientific, USA) coated with 300 ng/mL ACE2 and incubated for 60 minutes. The ACE2 was produced in HEK cells with a HAVT20 leader peptide, 10xhis-tag and a BirA-tag as described by Dekkers *et al*., [28]. After washing 5 times with PBS supplemented with 0.02% polysorbate-20 (PBS-T), plates were incubated for 30 minutes with streptavidin-poly-HRP (M2032, Sanquin). Plates were washed five times with PBS-T, and 100 μL of TMB substrate (100 μg/mL) and 0.003% (v/v) hydrogen peroxide (Merck, Germany) in 0.11 M sodium acetate buffer (pH 5.5) was added to each well. A total of 100 μL of 0.2 M H2SO4 (Merck, Germany) was added to stop the reaction. Absorbance was measured at 450 nm and 540 nm. The difference was used to evaluate RBD binding. As such, results were corrected for background signals (absence of RBD) and expressed as percentage binding relative to the uninhibited condition (% non-inhibited signal).

### Neutralization assay

Sera were tested by a SARS-CoV-2–specific virus neutralization test (VNT) based on a protocol described previously with some modifications [29]. In brief, replicate serial dilutions of heat-inactivated samples (30 minutes at 56°C) were incubated with 100-fold tissue culture 50% infectious dose of SARS-CoV-2 for 1 hour at 35°C. African green monkey (Vero-E6) cells were added in a concentration of 2 × 10^4^ cells per well and incubated for 3 days at 35°C in an incubator with 5% carbon dioxide. The 50% virus neutralization titer (VNT50), defined as the highest serum dilution that protected more than 50% of cells from cytopathological (lysed cells) effect, was taken as the neutralization titer. Samples with titers ≥10 were defined as SARS-CoV-2 seropositive.

### Data processing and analysis

Initial evaluation of IgG levels resulted in identification of 20 samples for which we observed either a >4-fold drop in level/wk or 4-fold difference/2 wk with consecutive follow-up before and after. These were excluded from all further analyses.

For the remaining 676 samples collected between March 30 and August 14, the longitudinal changes in IgG antibody levels were analyzed by linear mixed effects modeling in R (v3.6.0) using the LmerTest package (v3.1.2). We assumed first-order decay in concentrations, and therefore a linear trend in the log-transformed concentrations with time. Time was used as fixed variable, with random intercept for subject and random slope for time to account for individual clearance rates.

Statistical analysis were carried out using Graphpad Prism 7. Subgroup analysis on individual baseline parameters (Table 1) was performed using either a Mann-Whitney test or Z test for proportions. Correlations were evaluated as Spearman rank-order coefficient.

## Results

A total of 151 adult CCP donors were selected who were tested RT-PCR positive (nasopharyngeal swab) for SARS-CoV-2 (baseline information can be found in Table 1). On average, the first donation was collected 59 days after onset of COVID-19 symptoms. Additional plasma samples per donor were sequentially collected for up to 34 weeks after onset of symptoms, resulting in 844 plasma samples in total. Of these, 20 samples were excluded as outliers (details see M&M), resulting in 676 samples for **period 1** (up to 22 weeks after symptom onset; last sample median 110 days after symptom onset) and another 148 samples for **period 2** (extending to 34 weeks after symptom onset). During period 2, many donors were no longer donating due to insufficient titers, and this extended range is thus only available for a biased selection of plasma donors (n=55). Initial analysis of the antibody response to SARS-CoV-2 was therefore focused on period 1.

### Seroprevalence of antibodies against RBD

To study seroprevalence to SARS-CoV-2 we used our recently described sensitive total antibody bridging assay (so called RBD-Ab) to RBD [9]. All donors were positive for SARS-CoV-2 antibodies as assessed with the RBD-Ab assay, except for two samples, which belong to one donor that seroreverted at week 13 after tested positive at week 10 (Figure 1A). In addition, using isotype-specific assays [9] we determined seroprevalence of IgA, IgG and IgM to RBD. All samples collected before week 5 were seropositive for IgG against RBD (Figure 1A). As time progressed only a small fraction of samples fell below the detection limit for IgG, indicative that IgG levels remain in circulation for substantial periods after recovery, consistent with other studies [18, 19]. In contrast, the amount of samples that were seropositive in the IgA and IgM isotype specific assay were 75% and 68.8% before week 5, respectively, and gradually declined during the study period, in line with previous studies [18, 19].

**Figure 1.**
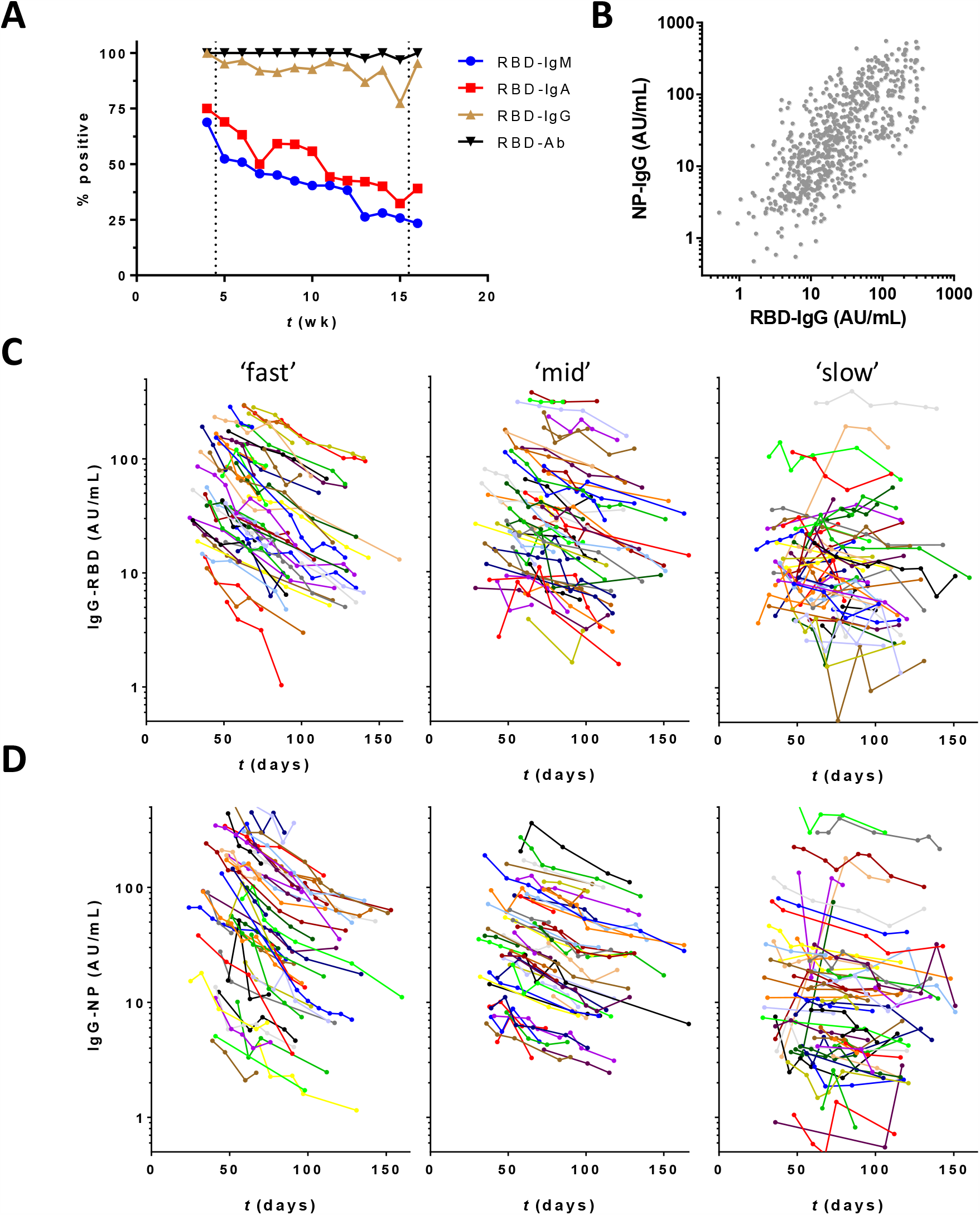
Antibodies against SARS-CoV-2 in CCP donors during up to 22 weeks of follow-up (period 1) (**A**) Seropositivity was assessed by isotype-specific (IgM, IgA and IgG) assays and the total antibody assay RBD-Ab. Results were stratified per week (wk) post onset symptoms; samples covering < 5 and > 15 weeks were combined. B) Correlation between anti-RBD and anti-NP IgG levels. Samples from the CCP donors were tested in the RBD and NP IgG isotype specific assay and their correlation was evaluated by Spearman (r = 0.73, p < 0.001). (**C**) Concentrations of IgG anti-RBD and (**D**) anti-NP plotted in days after onset of disease symptoms (676 samples from 151 donors). Left, middle and right panels contain samples that were stratified according to fitted half-lifes of antibody levels (see Figure 2).

### Levels of IgG antibody responses against RBD and NP

To obtain better insight into the antibody response we quantitatively analyzed the SARS-CoV-2 IgG antibodies against NP and RBD in all 151 adult donors. For (relative) quantification, we made use of pooled plasma of CCP donors as calibrator. Antibody concentrations varied >100-fold between individuals for both anti-RBD and anti-NP, in line with a recent study by Dan *et al*., [20] and were significantly correlated (Figure 1B). In addition, consistent with other studies high concentrations were significantly associated with hospitalization (Table 1) [30, 31]. Levels were also significantly higher in males than females. However, this is largely due to more men being hospitalized (22 vs 0%); and no significant difference was observed between non-hospitalized males and females (22 vs 17 AU/ml, and 22 vs 16 AU/ml, for anti-RBD and anti-NP, resp.; p = 0.28, and 0.14).

We also measured the IgG1 and IgG3 subclass responses to RBD at first donation in subclass-specific ELISA assays. To properly allow quantitative comparison, we recombinantly expressed IgG1 and IgG3 monoclonal antibodies [15], which were used as a calibrator. We observed that IgG anti-RBD consisted mainly of IgG1, median for IgG3 is 2.5% (IQR 1.5 – 5.1%; Figure S2A). Since higher subclass ratios were reported for antibodies against the S2 vs S1 domain of the S protein [32, 33], we also tested IgG1 and IgG3 in a subset of samples against the full S protein, but we obtained very similar results (Figure S2B).

Since titrations of the recombinant antibodies were parallel to the plasma pool used as calibrator (Figure S2D), this also enabled us to estimate the absolute concentration of RBD-reactive IgG antibodies. The average concentration of anti-RBD IgG observed in CP donors of 24 AU/ml was found to reflect approximately 2-3 µg/mL.

### Dynamic changes in IgG against SARS-CoV-2

Next, we assessed the dynamics of the IgG antibody response over time. Within the time window of up to 22 weeks, most individuals demonstrated a steady, and approximately log-linear decline in antibody concentration (Figure 1C,D). Assuming a log-linear (i.e., first-order) decline in antibody concentrations, we analyzed this trend using linear mixed effects modeling, and found a median half-life of 62 (IQR 48 – 87) and 59 (IQR 44 – 82) days for anti-RBD and anti-NP IgG levels, respectively (Figure 2A and 2B and Table 1). There is a very weak correlation between estimated half-life for anti-RBD IgG and anti-NP IgG levels, as shown in Figure 2C (Spearman r = 0.23, p = 0.0037). However, the subset of individuals demonstrating the most rapid decline do so for both types of antibodies. There is a significant but weak negative correlation between half-life and absolute levels (Figure S1), indicating a more rapid decline in individuals with the highest antibody levels.

**Figure 2.**
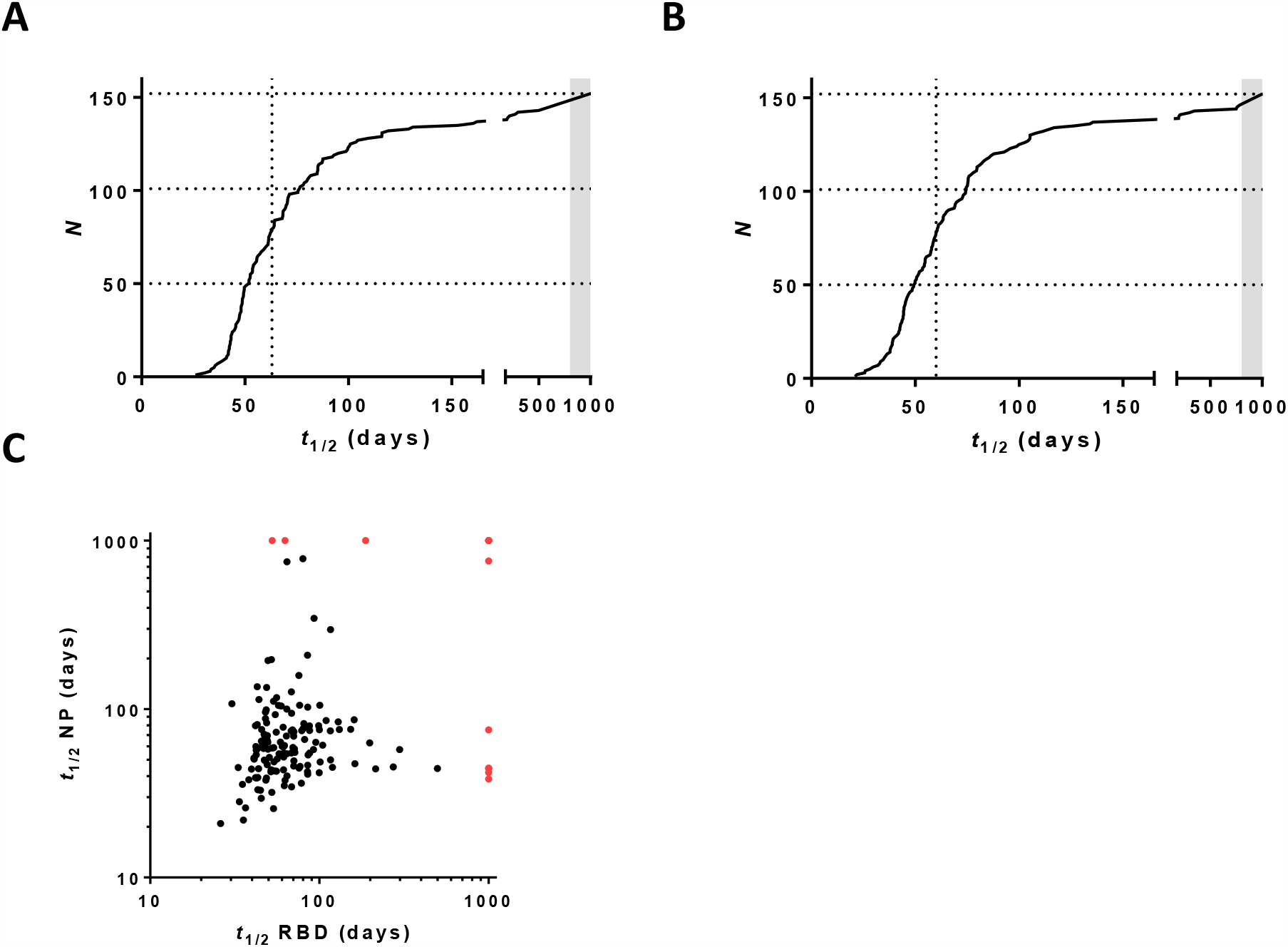
Regression analysis of IgG levels. For both (**A**) RBD-IgG and (**B**) NP-IgG data of period 1 was modeled using a mixed effects model (log-linear in IgG; random intercept and slope, time as fixed variable). Slopes of IgG decay in time (see Figure 1) were converted into half-lifes. Dotted vertical lines indicate median half-lifes. (**C**) The correlation between estimated half-lifes for anti-RBD IgG and anti-NP IgG levels was evaluated by Spearman (r = 0.23, p = 0.0037). In case of rising levels, *t* was arbitrarily assigned a value of 1000 in the above figures (indicated by grey bar (**A**,**B**) and red dots (**C**), respectively).

Although IgG3 has a shorter half life than IgG1, we did not observe an association between IgG clearance rate and IgG1 and IgG3 levels in the first sample (Figure S2C), also not for those samples with a relatively high percentage of IgG3 antibodies. In fact, the half-lifes of the IgG anti-RBD and anti-NP levels exceed the intrinsic half-life of both IgG3 antibodies (ca. 7 days) and IgG1 antibodies (ca. 21 days) [34], in line with continued antibody production.

For a subset of 55 donors extended follow-up was available for up to 34 weeks (period 2). Of note, as explained in the M&M, there is a selection bias in this group towards higher antibody levels for period 2. Overall, the declining trends in antibody levels continued (Figure 3A,B). Interestingly, the rate of decline appears to decrease at later time points, especially for donors with an initially high decline (Figure S3). The effect is more pronounced for anti-RBD (Figure 3C) than anti-NP (Figure 3D).

**Figure 3.**
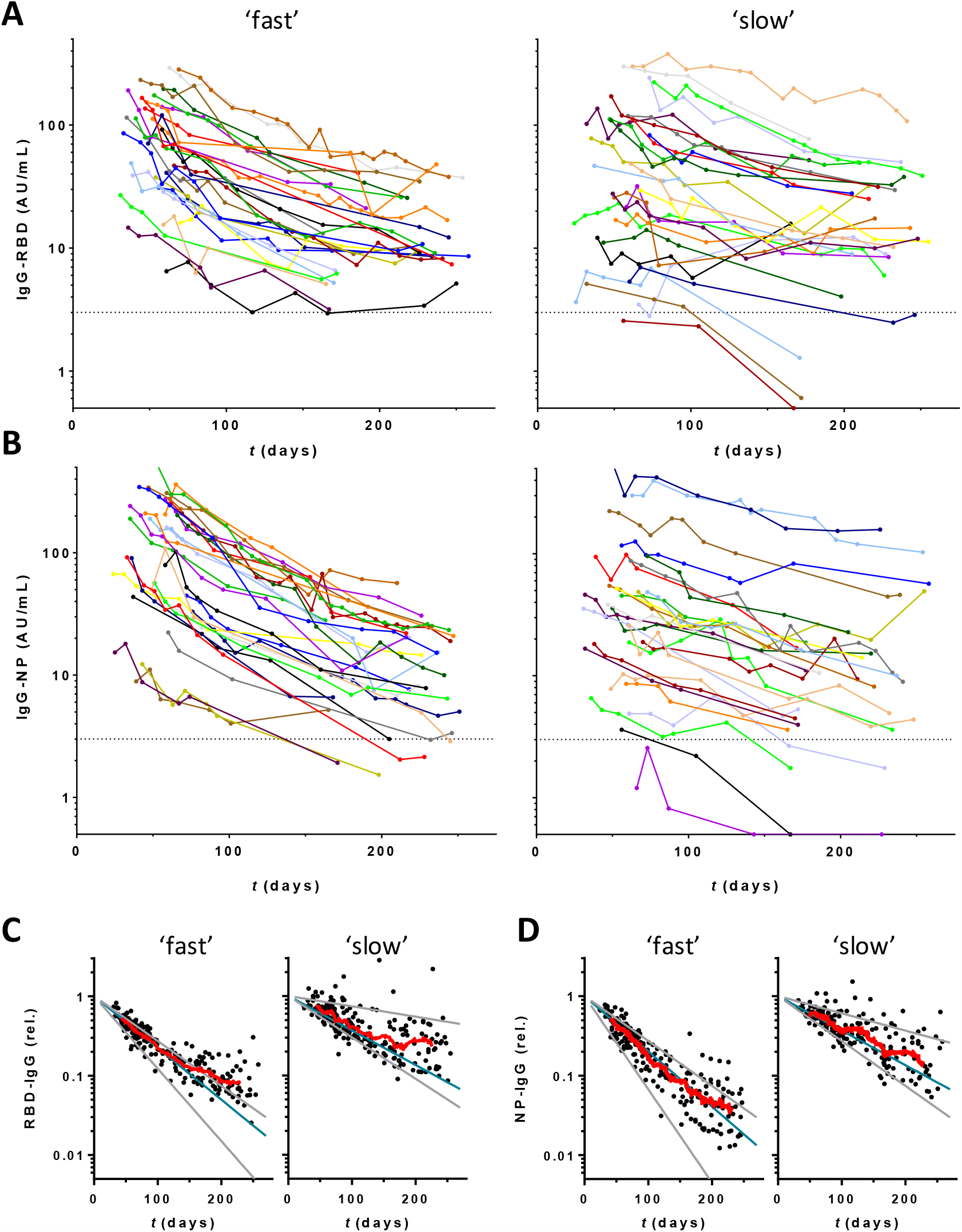
Dynamic changes in IgG antibodies against SARS-CoV-2 during extended follow-up up to 34 weeks (period 2) Concentrations of (**A**) IgG anti-RBD and (**B**) anti-NP plotted in days after onset of disease symptoms (430 samples from 55 donors). Left panels 28 donors with fastest decline during period 1, right panels slowest 27 donors. (**C**,**D**) Same data but normalized per donor using fitted intercepts from regression analysis of period 1. Blue and grey lines indicate median, smallest, and largest fitted slopes from same analysis (excluding positive slopes, 2 for RBD and 1 for NP) within both groups. Red lines are running averages showing overall trend within both groups of donors.

### IgG levels correlate with viral neutralization

To obtain insight into the neutralizing capacity of the antibody response in this population, we carried out a competition ELISA in which binding of RBD to ACE2, the receptor on SARS-CoV-2 target cells, is inhibited by blocking antibodies from plasma samples (Figure S4). We observed a high and significant correlation between anti-RBD IgG levels and the amount of competition (Figure 4A), independent of the timing of plasma collection (Figure 4B). On the other hand, correlation between anti-RBD IgM or IgA and competition was very weak (Figure S5). This indicates that IgG probably plays an important role in neutralizing SARS-CoV-2 virus particles, especially at the long term. Furthermore, we tested virus neutralization of a subset of the plasma samples (n = 147) using the well-established classic plaque reduction assay using live SARS-CoV-2 virus. We observed a good correlation between plaque reduction, expressed as the titer that reduced plaque formation by 50% (VNT50), with anti-RBD IgG (Figure 4C) as well with viral neutralization (Figure 4D) in the in-house competition assay. This data also indicates that our ELISA-based competition assay could be used to evaluate the virus neutralizing capacity of anti-SARS-CoV-2 antibodies in plasma samples.

**Figure 4.**
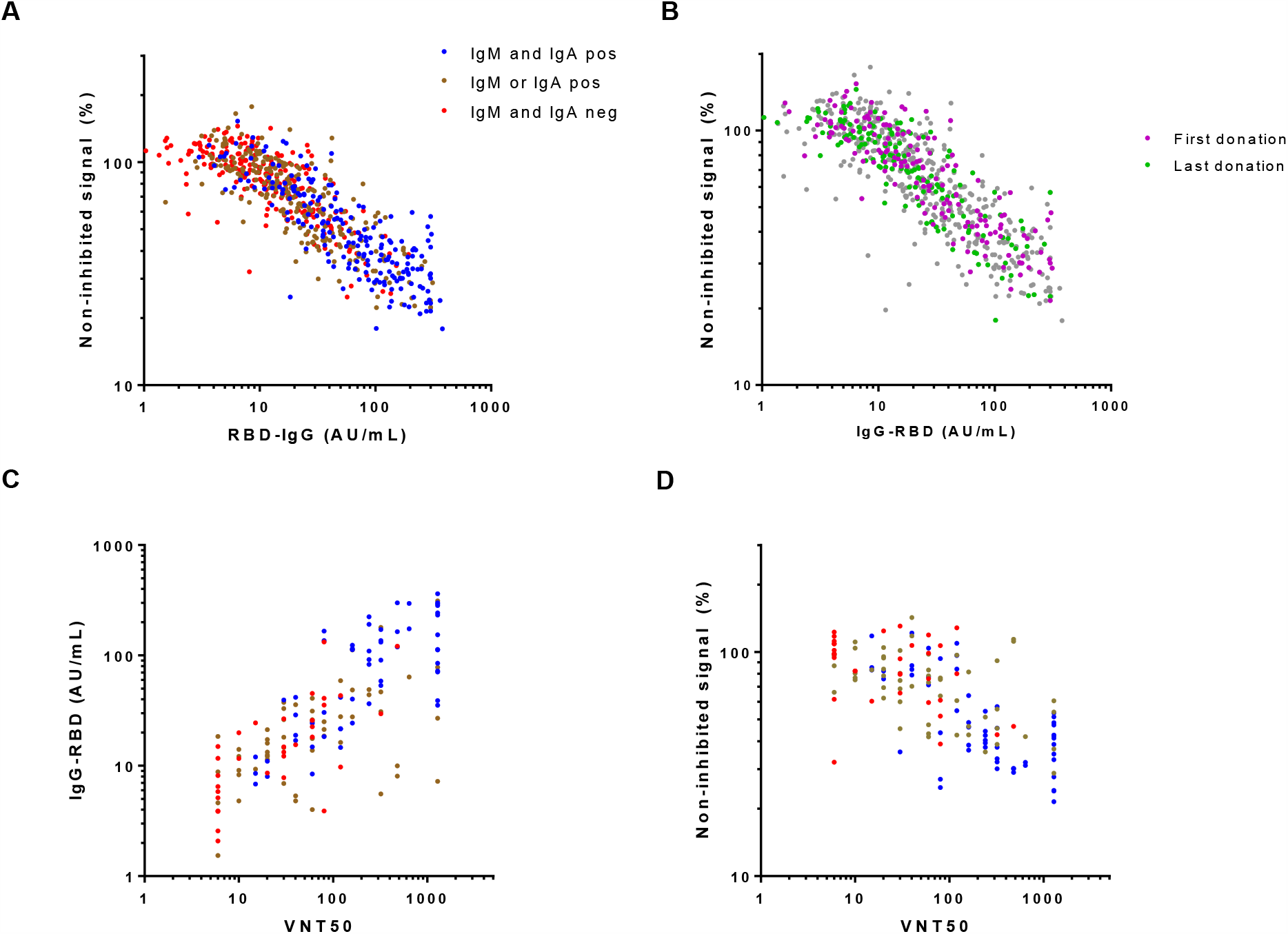
Correlation between IgG levels and virus neutralization. Plasma samples were tested in the in-house developed competition ELISA (676 samples from 151 individual donors), and in the classic plaque reduction assay (147 samples from 129 individual donors). The correlation between anti-RBD IgG and virus neutralization in the (**A, B**) competition assay and (**C**) plaque reduction assay was assessed by Spearman (r = 0.85, r = 0.75, resp., p < 0.001). (**D**) A correlation between the two viral neutralization assays was also observed (Spearman r = 0.65, p < 0.001).

## Discussion

Here we describe the individual dynamics of the longitudinal antibody response in plasma samples from CCP donors that were followed for up to 34 weeks post onset of COVID-19 disease symptoms. IgG levels against RBD and NP decreased only gradually, albeit with substantial interindividual variation, and the decrease appeared to slow down as time progressed. This information is relevant not only in the context of the persistence of detectable antibody responses over time and the assessment of seroprevalence and immunity in populations, but may also be used to improve the selection procedure of CCP donors for therapies with convalescent plasma.

The median half-lifes of 62 (48 – 87) and 59 (44 – 82) days for anti-RBD and anti-NP IgG, respectively, are reasonably similar if slightly shorter to those found in Dan *et al*., [20], that reported a half-life of 83 days (62 – 127) for anti-RBD IgG and 67 days (49 – 105) for anti-NP IgG. The difference may be explained by the fact that Dan *et al*. only used paired samples from individuals that donated 6 days and 8 months post onset of symptoms. Our study included a median number of 5 samples per donor, which allowed a more thorough analysis of individual rate profiles. The data suggests that the decline in antibody levels is not uniform during this 8 months period, but continues at a slower pace at later time points. The reported half-lifes in this study reflect the period between ca. 2 and 5 months post symptom onset. Interestingly, individuals showed a considerable, several-fold differences in IgG half-lifes. A limitation is the substantial uncertainty in the individual half-life estimates especially at the low end, since the available data will sometimes only cover a fraction of a half-life.

Long-term maintenance of antibody production is provided by a pool of long-lived plasma cells and memory B-cells that can last a lifetime [35]. Several time scales may be identified, of which the longest-lasting can have half-lifes in the order of many years, and thus beyond scope of the current study [36]. It will be interesting to continue following the decline of anti-RBD IgG in time and investigate whether the longevity of the antibody response is similar to other coronaviruses, such as SARS-CoV that can still be detected in most individuals 3-years after recovery [37]. The antibody titer required for protection against re-infection in humans is not yet known, and has to be evaluated also in the context of a recall response upon re-infection. Recent studies provide evidence that upon infection, individuals develop SARS-CoV-2 specific memory B and memory T cells that can be detected for up to 240 days, with numbers of IgG memory B cells increasing over time and plateauing after ca. 150 days [20, 38]. The relationship between long-lasting antibody production and the T and B cell memory compartments requires more investigation. We also quantitatively examined the IgG1 and IgG3 subclass response and found that in our study population IgG1 appeared to be by far the dominant IgG subtype. This is in contrast to previous studies that suggest an IgG subclass ratio skewed towards IgG3 during SARS-CoV-2 infection [33, 39]. A possible explanation for this discrepancy is that in these studies the plasma samples were collected shortly after COVID-19 disease recovery, while in our study samples were collected up to 22 weeks after onset of disease symptoms. A strength of the current study is the use of matching IgG1 and IgG3 monoclonal anti-S antibodies that allowed reliable quantification of the relative amounts of IgG1 and IgG3.

The dynamics of the SARS-CoV-2 antibodies play an important role in CCP donor recruitment in order to select the most optimal timing of plasma collection, in which the level of IgG probably plays a crucial role. Potent CCP units should in theory contain high amounts of neutralizing antibodies against SARS-CoV-2. We observed a good correlation between anti-RBD IgG and both a classic plaque reduction viral neutralization assay, consistent with previous studies [40, 41], as well as a straightforward competition ELISA that indirectly assesses viral neutralization by measuring the ability of plasma containing anti-SARS-CoV-2 antibodies to prevent interaction between RBD and ACE2. This suggests that a competition assay may also reliably report on viral neutralization. Of note, a recent study by Gasser *et al*., found that depletion of IgM resulted in a substantial loss of virus neutralization of the corresponding plasma samples, suggesting a role of IgM in virus neutralization [42]. However, IgM (and IgA) levels will drop relatively fast after recovery. Indeed, many plasma donations did not contain detectable amounts of IgM or IgA. Accordingly, we observed only a very weak correlation between viral neutralization in the competition assay and IgM levels.

In line with other studies, higher IgG levels were found for patients that were hospitalized than those with milder symptoms not requiring hospitalization, indicating that severely ill patients are more suitable for CCP donation [21]. Nevertheless, there are potential caveats for these donors. First, a recent study by Larsen *et al*., showed that severely ill COVID-19 patients with acute respiratory distress syndrome may display a so-called “afucosylated IgG anti-S response” [43, 44]. In addition, multiple studies reported the presence of autoantibodies against a.o. type I IFN-α2 and IFN-ω in patients with severe COVID-19 pneumonia that also show high IgG titers [45-49]. These parameters might negatively impact the outcome of therapies with convalescent plasma, and suggest that it may be safer to rely on convalescent plasmas from patients with mild symptoms, despite the fact that those tend to have lower antibody-response. In conclusion, this study provides insight into the individual dynamics of antibody levels to SARS-CoV-2 during up to 34 weeks after symptom onset. Substantial (several-fold) variation in individual half-lifes was observed, with median half-lifes of around 60 days for both anti-RBD and anti-NP IgG, and a tendency towards slower rates of decline of antibody levels as time progressed.

## Data Availability

The data is available upon reasonable request (by contacting the corresponding author).

## Acknowledgements

We would like to thank all the donors who have kindly donated plasma.

## List of abbreviations

RBD: receptor binding domain
NP: nucleocapsid protein
S: spike
SARS-CoV-2: severe acute respiratory syndrome coronavirus 2
ACE2: angiotension converting enzyme-2
VNT50: virus neutralization titer 50%
Ab: antibody
PTG: PBS supplemented with 0.02% polysorbate-20 and 0.3% gelatin

## Supplementary information

**Figure S1.**
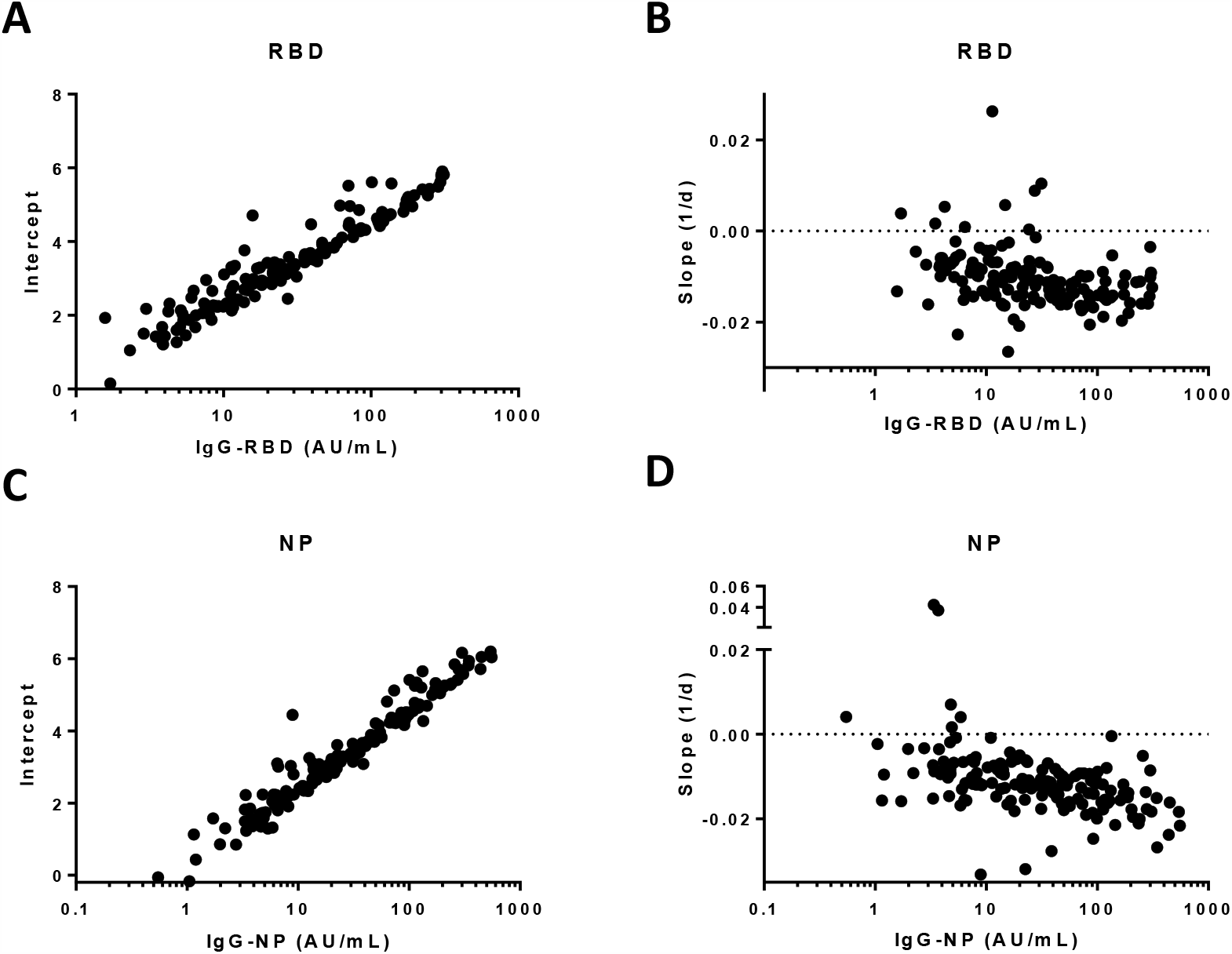
Correlation plots. Plots for fitted intercepts and slopes for (**A, B**) RBD-IgG and (**C**,**D**) NP-IgG with respect to measured IgG levels at first donation. Interaction between slope and intercept/baseline IgG level: r = 0.22, p = 0.006 (RBD) and r = 0.34, p < 0.0001 (NP).

**Figure S2.**
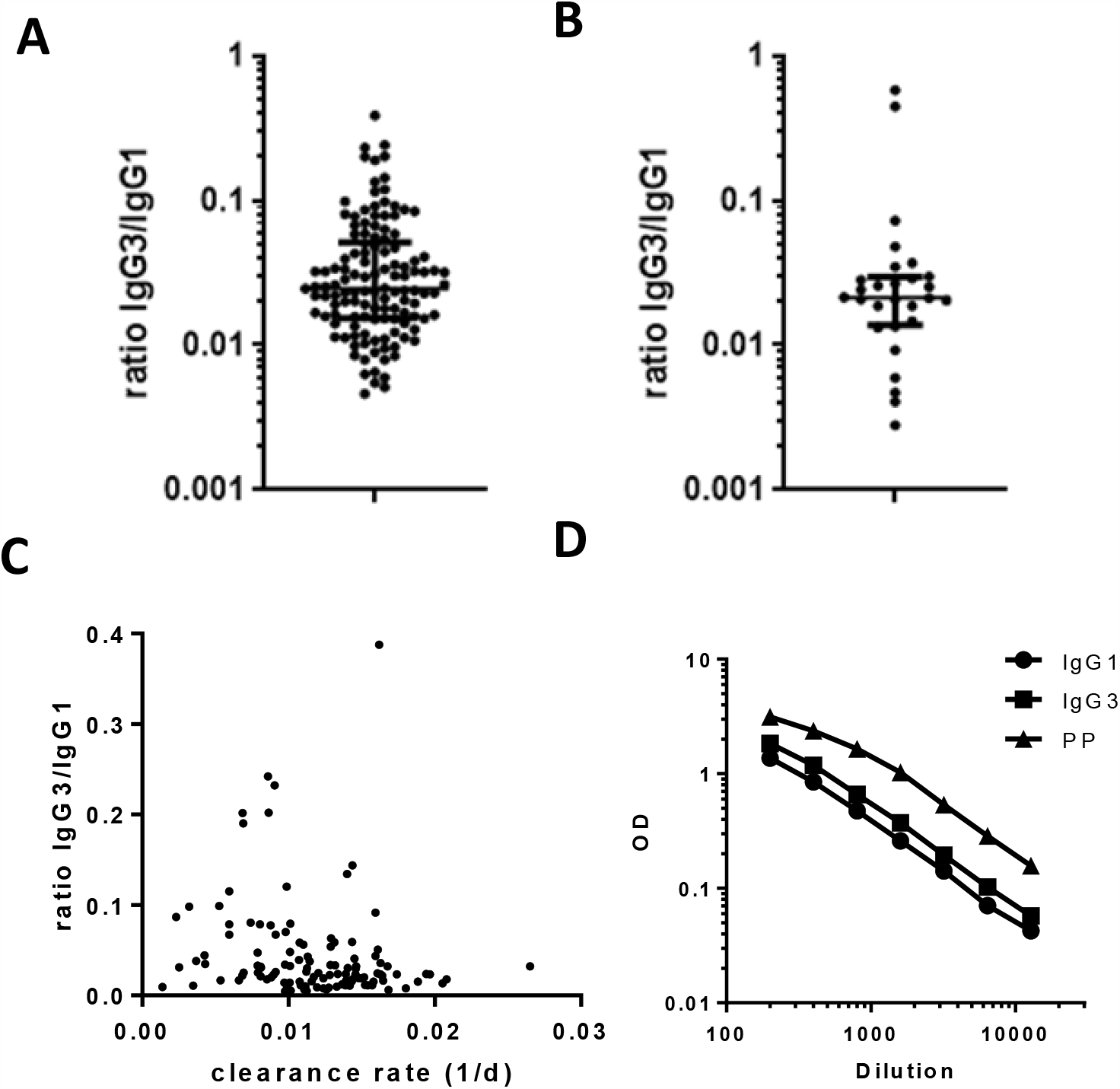
Contribution of IgG1 and IgG3 subclasses to anti-RBD. IgG1 and IgG3 antibody subtypes were measured against (**A**) RBD and (**B**) S1 and displayed as ratio IgG3 compared to IgG1. For RBD, first available sample for each donor was measured, for S, a random selection of 30 donors was selected. (**C**) IgG3/IgG1 ratio vs clearance rate. (**D**) Titrations of IgG1 and IgG3 versions of the monoclonal antibody 1-18, and the plasma pool (PP) that was used as calibrator in this study.

**Figure S3.**
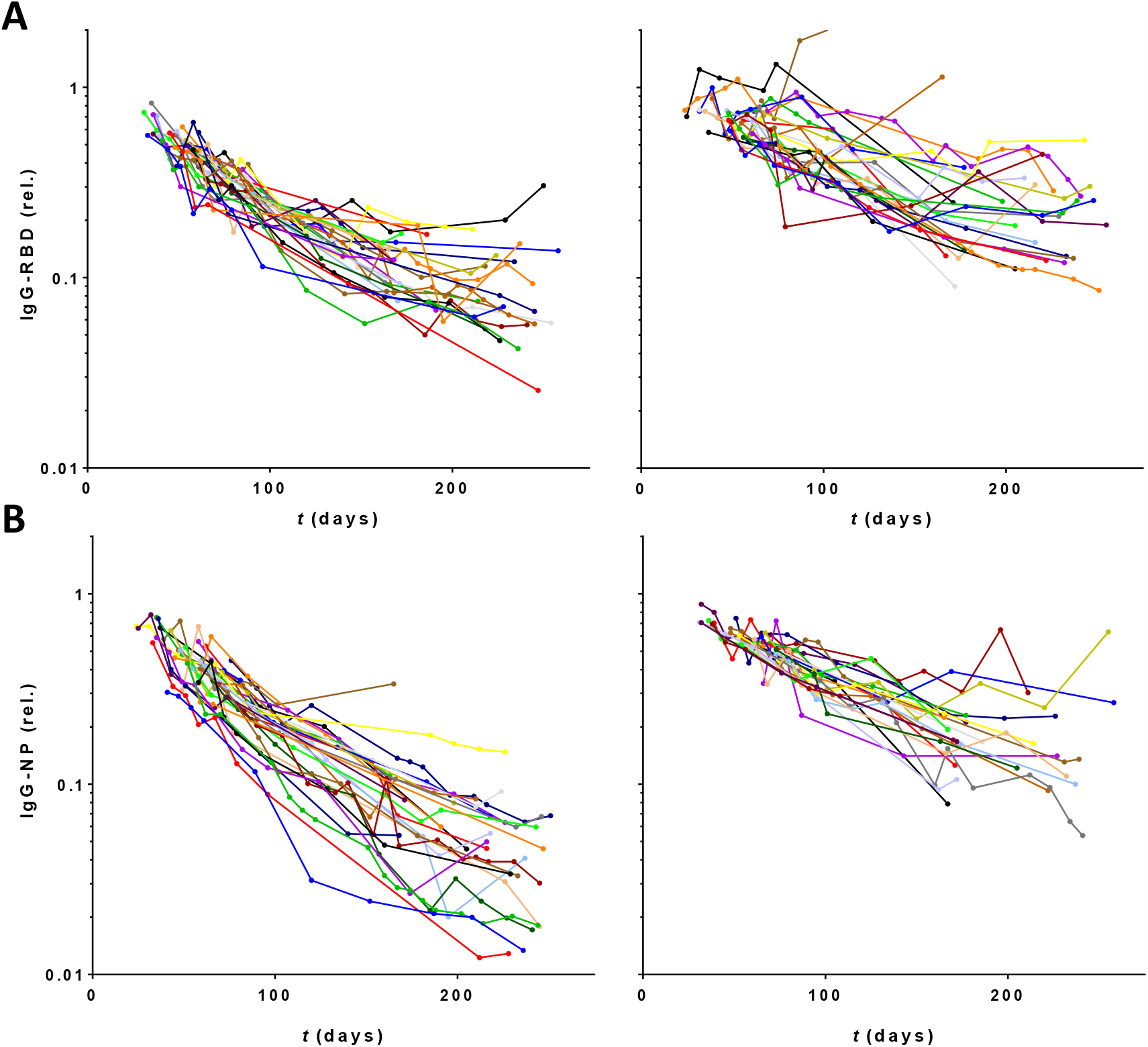
Normalized individual trends in IgG antibody levels during extended follow-up up to 34 weeks (period 2) Concentrations of (**A**) IgG anti-RBD and (**B**) anti-NP, normalized per donor using fitted intercepts from regression analysis (Figure 2), plotted in days after onset of disease symptoms (430 samples from 55 donors). Left panels 28 donors with fastest decline during first 20 weeks, right panels slowest 27 donors.

**Figure S4.**
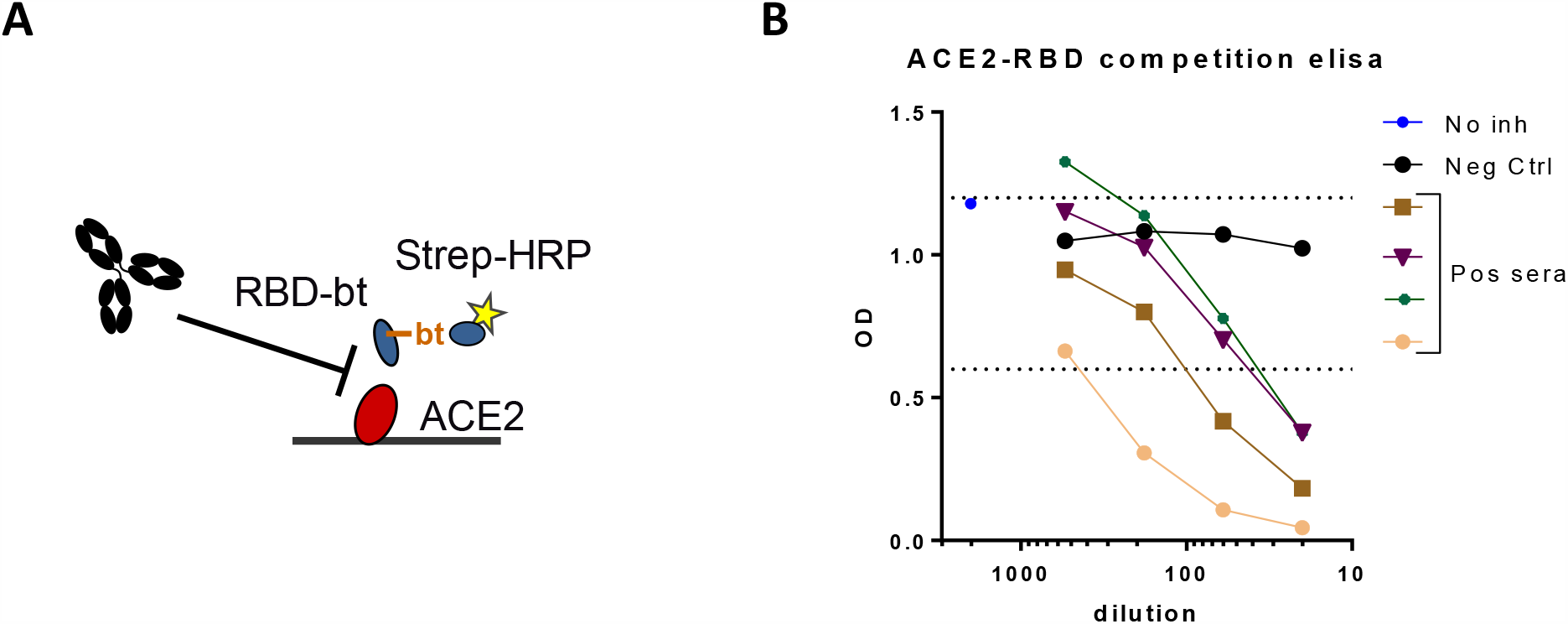
Overview of the competition assay. (**A**) schematic representation of the assay format. Biotinylated RBD will bind to immobilized ACE2 only to the extent that neutralizing antibodies from serum do not inhibit this interaction. (**B**) dose-response curves for several sera containing antibodies to Sars-CoV-2.

**Figure S5.**
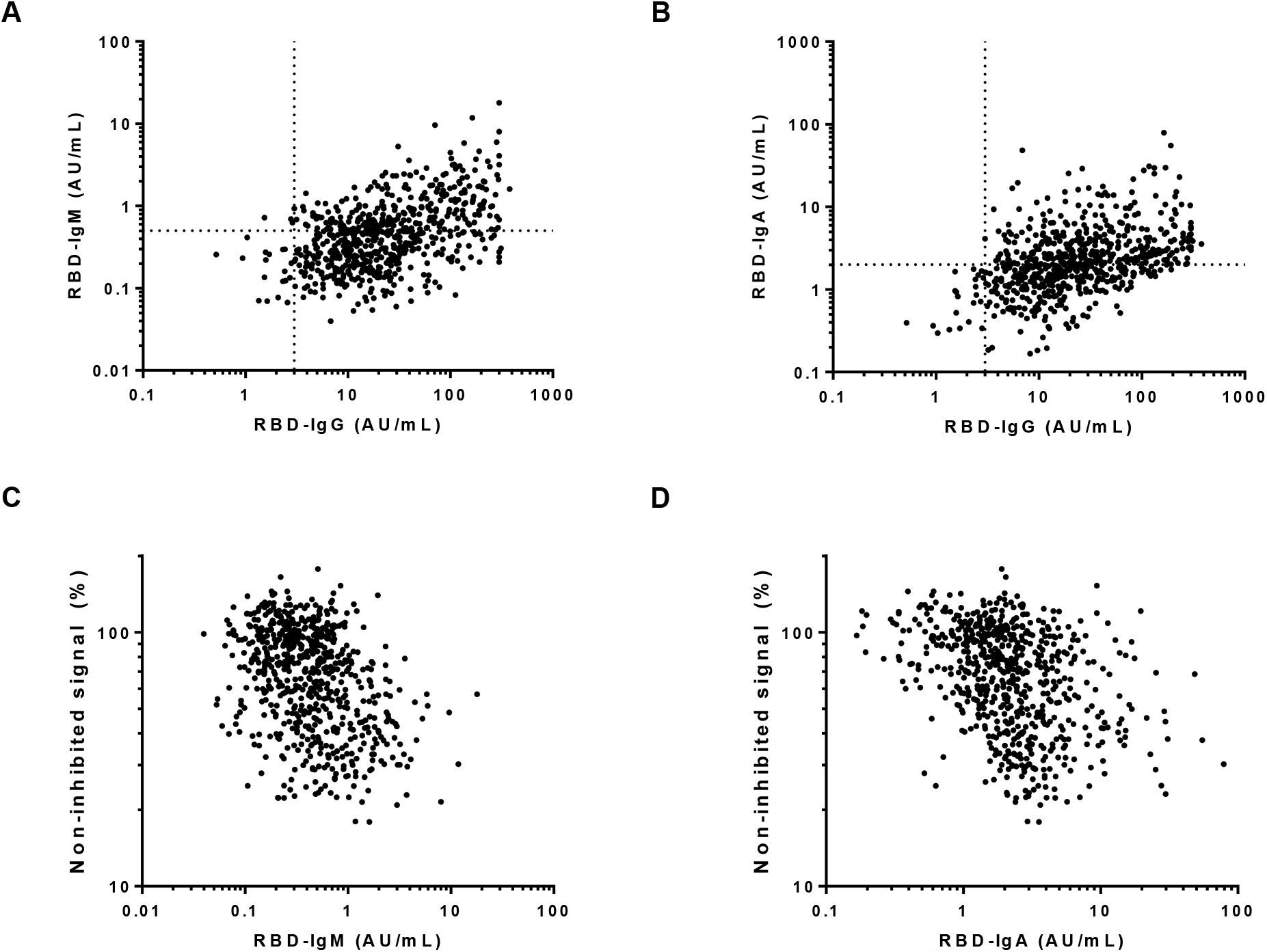
Correlations of IgM and IgA anti-RBD with IgG anti-RBD and ACE2-RBD competition. Correlation between IgM (**A**) and IgA (**B**) with IgG anti-RBD in 676 samples of 151 donors collected up to week 20 after symptom onset. Spearman r = 0.41 (p < 0.0001), and 0.23 (p < 0.001), resp. Correlation between competition ELISA and IgM (**C**) and IgA (**D**) in the same set of samples. Spearman r = −0.26 (p < 0.001), r = 0.22 (p < 0.001), resp.

